# The left inferior frontal gyrus is causally involved in selective semantic retrieval: Evidence from tDCS in primary progressive aphasia

**DOI:** 10.1101/2020.07.20.20151043

**Authors:** Zeyi Wang, Bronte N Ficek, Kimberly T Webster, Chiadi U Onyike, John E Desmond, Argye E Hillis, Constantine E Frangakis, Caffo Brian, Tsapkini Kyrana

**Author notes:** Corresponding author: Kyrana Tsapkini, Ph.D., Department of Neurology, 600 N. Wolfe Street, Phipps 488, Baltimore, MD 21287, USA.

## Abstract

Lesion and imaging studies have shown that the left inferior frontal gyrus (IFG) is involved in selective semantic retrieval of information from the temporal lobes. However, causal, i.e., interventional, evidence is sparse. In the present study we addressed this question by testing whether transcranial direct current stimulation (tDCS) over the left IFG in a group of individuals with primary progressive aphasia may improve semantic fluency, a task that relies to selective semantic retrieval. Semantic fluency improved significantly more in the tDCS vs. sham condition immediately post-treatment and improvement lasted up to 2 months. We further addressed the question of who will benefit most from such an intervention by testing possible demographic, clinical and functional connectivity variables that may predict the behavioral tDCS effect. We found that patients with stronger baseline functional connectivity between the subareas of the left IFG opercularis and triangularis, and between the middle temporal pole and superior temporal gyrus. were the most likely to benefit from tDCS over the left IFG. We thus provided causal evidence that the left IFG is the neural substrate of selective semantic retrieval and tDCS over the left IFG may improve semantic fluency in individuals with stronger baseline functional connectivity.

## 1. Introduction

Semantic fluency is a neuropsychological task in which a participant is asked to generate words that belong to specific semantic categories, such as fruits or animals (Lezak et al. 2004). The task of semantic fluency relies on the function of selective semantic retrieval. The distinction between function and task was at the heart of cognitive psychology and neuropsychology from the time of box diagrams (Caramazza 1997). Interestingly, from the early days of functional neuroimaging, it became evident that most brain areas, although they may specialize in certain functions (computations), are usually involved in several tasks (for a review, see Price, 2012). This understanding allows for the exciting possibility that if we were able to modulate the neural function of a particular area, then we would potentially modulate the cognitive function (computation) it performs and, consequently, several tasks that involve this function.

Functional magnetic resonance imaging (fMRI) studies in healthy controls have shown that selective semantic retrieval is regulated by the left inferior frontal gyrus (IFG) triangularis (Brodmann’s area [BA] 45/47), which retrieves semantic information stored in the temporal brain regions (Binder & Desai, 2011; Petrides, 2006; Rolheiser, Stamatakis, & Tyler, 2011; Thompson-Schill, D’Esposito, Aguirre, & Farah, 1997; Tyler et al., 2011). Further fMRI and diffusion tensor imaging (DTI) studies have shown that the left IFG triangularis is directly and monosynaptically connected to the temporal lobes via the extreme capsule fasciculus and that these pathways are critical for language comprehension (Frey et al. 2008; Saur, Kreher, Schnell, Kümmerer, Kellmeyer, Vry, Umarova, Musso, Glauche, Abel, et al. 2008; Rolheiser et al. 2011b). However, causal inference for the role of a particular brain area and a particular computation comes only from direct intervention studies targeting this area, such as in vivo electrical stimulation mostly during epilepsy surgery (see Rofes et al. 2018 for a review on language functions) or, more recently, neuromodulation studies such as transcranial direct current stimulation (tDCS).

Anodal tDCS is believed to induce long-term potentiation (LTP) in the stimulated brain region(s) by raising neuronal resting membrane potentials and thereby increasing excitability; however, the exact mechanisms are still an area of active research (Nitsche and Paulus 2000, 2011; Schlaug et al. 2011). TDCS has been shown to benefit language performance in post-stroke aphasia (Monti et al. 2008; Baker et al. 2010; Chrysikou and Hamilton 2011; Fiori et al. 2011; Fridriksson et al. 2011; Kang et al. 2011; Marangolo et al. 2011), and more recently as a potential adjunct to therapy in primary progressive aphasia (Mesulam, 2001, 2003, 2008), a neurodegenerative syndrome in which language abilities gradually deteriorate while other cognitive functions remain relatively intact in the early years of the condition (Cotelli et al. 2014; Tippett et al. 2015; Tsapkini et al. 2018). In the tDCS literature, many studies show transfer to untrained items/words. For example, in our recent clinical trial in 36 PPA participants we found variant effects in transfer to untrained items (Tsapkini et al. 2018; Cotelli et al. 2019). Few studies, however, evaluate generalization of tDCS effects to untrained tasks even if they subserve the same function: two in post-stroke aphasia (Marangolo et al. 2011; Meinzer et al. 2014) and 3 in PPA (Cotelli et al. 2014; Gervits et al. 2015; Roncero et al. 2017). From a clinical perspective, generalization of improvement is the most desirable outcome of an intervention approach, particularly because syndromes, such as PPA, often affect several language functions and performance deteriorates with time.

Given the small number of tDCS studies in PPA that show transfer effects to untrained tasks (only 3), we refer to each one separately. Cotelli and colleagues (2014) have shown that anodal tDCS over the left dorsolateral prefrontal cortex improved naming accuracy in 16 patients with non-fluent variant PPA (nfvPPA), and significant improvement was also found in self-assessments of speech production on functional communication scales (Cotelli et al. 2014). In another recent study, Gervits and colleagues found that six people with PPA, who narrated wordless children’s books while undergoing 10 sessions of tDCS over the left frontal cortex (centered at F7), found significantly improved category (semantic) fluency compared to sham at follow-up intervals (Gervits et al. 2015). Roncero and colleagues (2017) also found transfer effects to untrained picture-naming items and digit span (forward and backward, [Wechsler 1981]) after tDCS over the left inferior parietal cortex in 10 patients with PPA (Roncero et al. 2017). Therefore, there is encouraging preliminary evidence of transfer to other tasks, especially from Gervits and colleagues’ study (i.e., transfer from a lexical retrieval task to semantic fluency). Furthermore, two additional studies directly targeted semantic fluency and found that a single session of tDCS over the left IFG improved semantic fluency in healthy controls showcasing its causal involvement in selective semantic retrieval (Cattaneo et al. 2011; Penolazzi et al. 2013).

Given the above evidence, we hypothesized that if the function of the left IFG (particularly the pars triangularis) is selective semantic retrieval of information stored in the temporal lobes (Thompson-Schill et al. 1997, 1999; Petrides 2006), then tDCS over the left IFG would also improve performance in other, non-trained tasks involving the function of active selective semantic retrieval, such as semantic fluency. In the present study, we used neuromodulation, a clinical intervention method, to address a neurocognitive question, i.e., whether the left IFG is causally involved in selective semantic retrieval, while at the same time addressing the important clinical question of treatment transfer to untrained tasks that rely on the same function (computation). The present study aimed to determine whether the tDCS effects in trained written-naming task can transfer to untrained semantic fluency tasks in a large cohort of patients with PPA—the largest PPA cohort, to our knowledge. Furthermore, in order to elucidate why transfer effects might occur and what are the characteristics of those individuals who may benefit from tDCS, we tested demographic, clinical and functional connectivity pairs as predictors of the semantic fluency modulation due to tDCS over the left IFG.

## 2. Materials and Methods

### 2.1. Participants

Thirty-six patients with PPA participated in this study (17 female): 14 with logopenic variant PPA (lvPPA), 13 with non-fluent variant PPA (nfvPPA), and 9 with semantic variant PPA (svPPA). All were right-handed, native English speakers, between 50 and 80 years old, and diagnosed based on clinical assessment, neuropsychological and language testing, and MRI, according to consensus criteria (Gorno-Tempini et al. 2011). Informed consent was obtained from participants or their spouses, and all data were acquired in compliance with the Johns Hopkins Hospital Institutional Review Board. Figure 1 shows the participants recruited and their randomization to tDCS or sham condition. Each PPA variant group was matched by sex, age, education, years post onset of symptoms, overall FTD-CDR score and language severity measures (Tables 1A, 1B).

**Table 1A.**
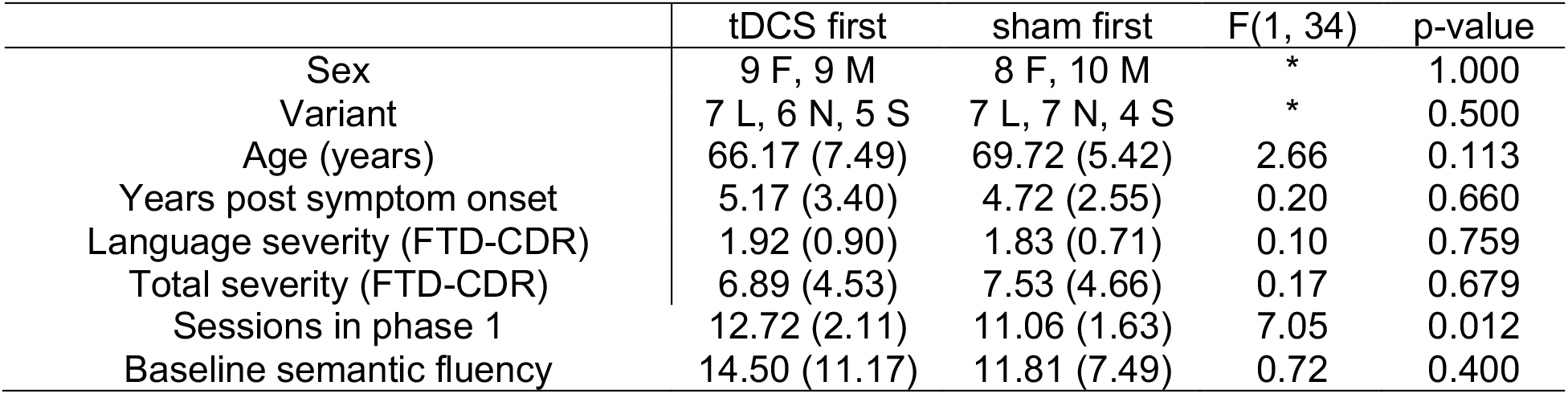
Means and standard deviations of demographic variables and baseline semantic fluency scores grouped by first-phase condition (n=36). *Fisher’s exact test used. FTD-CDR, Frontotemporal Dementia Clinical Dementia Rating Scale sum of boxes (Knopman et al. 2008). F, female; M, male. L, logopenic; N, nonfluent; S semantic.

**Table 1B.** Means and standard deviations of demographic variables and baseline semantic fluency scores grouped by PPA variant (n=36). *Fisher’s exact test used. FTD-CDR, Frontotemporal Dementia Clinical Rating Scale sum of boxes (Knopman et al. 2008). F, female; M, male. s, sham; t, tDCS.

|  | lvPPA | nfvPPA | svPPA | F(2,33) | p-value |
| --- | --- | --- | --- | --- | --- |
| Sex | 7 F, 7 M | 5 F, 8 M | 5 F, 4 M | * | 0.800 |
| First-phase condition | 7 s, 7 t | 7 s, 6 t | 4 s, 5 t | * | 1.000 |
| Age (years) | 66.29 (8.11) | 69.77 (6.00) | 67.89 (4.96) | 0.91 | 0.412 |
| Years post symptom onset | 4.82 (3.33) | 4.65 (2.66) | 5.56 (3.08) | 0.25 | 0.780 |
| Language severity (FTD-CDR) | 1.57 (0.83) | 2.04 (0.72) | 2.11 (0.78) | 1.76 | 0.188 |
| Total FTD-CDR | 6.18 (3.76) | 7.85 (4.19) | 7.89 (6.17) | 0.57 | 0.571 |
| Sessions in phase 1 | 11.93 (2.02) | 11.85 (1.91) | 11.89 (2.47) | 0.01 | 0.990 |
| Baseline semantic fluency | 17.50 (10.97) | 12.08 (8.38) | 7.94 (5.15) | 3.30 | 0.049 |

**Figure 1.**
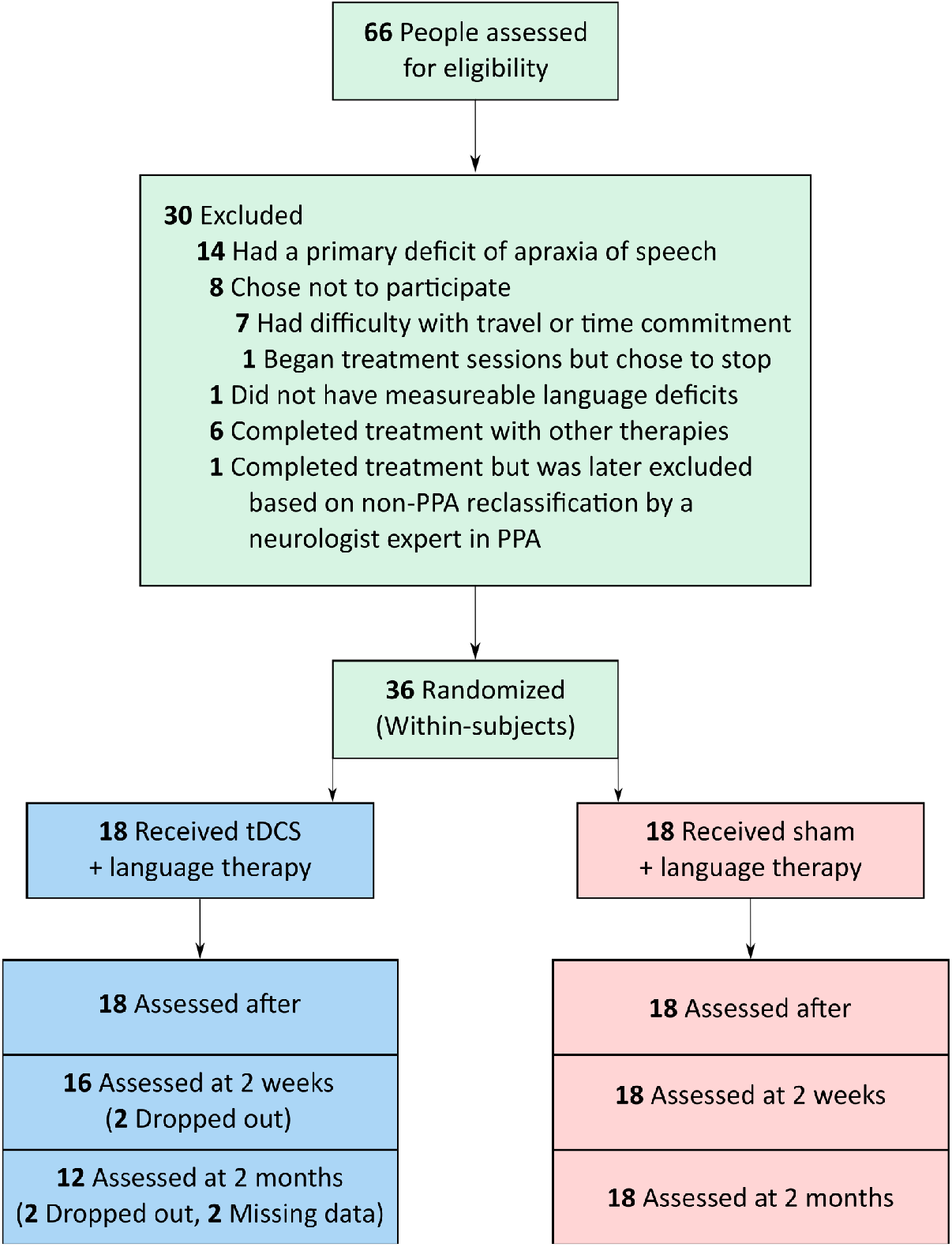
Participants recruited and randomization to tDCS or sham.

### 2.2. Overall design

We used a within-subjects, double-blind, crossover design with two experimental conditions: speech-language therapy plus conventional anodal tDCS over the left IFG, and speech-language therapy plus sham tDCS. Each condition lasted approximately 12 consecutive weekday sessions; the two phases were separated by a 2-month wash-out period. Evaluations—consisting of a set of trained and untrained items of the same task, as well as extensive neuropsychological and neurolinguistic assessments— occurred immediately before, immediately after, two weeks after, and two months after each treatment phase. Participants, speech-language therapists, and examiners were blind to the experimental condition.

### 2.3. tDCS methods

Each daily therapy session lasted one hour. For both active tDCS and sham conditions, two 5cm x 5 cm, non-metallic, conductive, rubber electrodes covered with saline-soaked sponges were placed over the right cheek (cathodal electrode) and the left IFG centered at F7 of the EEG 10-20 electrode position (anodal electrode) (Homan 1988). The electrodes were hooked up to a Soterix 1×1 Clinical Trials device, which elicited a tingling sensation on the scalp as it ramped up within 30 seconds, to deliver current at an intensity of 2 mA (estimated current density 0.08 mA/cm^2^; estimated total charge 0.096 C/cm^2^). In the tDCS condition, current was delivered for 20 minutes for a daily maximum of 2.4 Coulombs; in the sham condition, current ramped up to 2 mA over a 30 sec interval and immediately ramped down to elicit the same tingling sensation, a procedure that has been shown to blind participants to treatment condition (cite Gandiga 2006 paper). Stimulation started at the beginning of each therapy session and lasted for 20 min whereas speech-language therapy continued for a full session, i.e., 25 additional minutes, for a total of 45-50 min of a regular speech-language therapy session. Twice during each session, participants rated their level of pain with the Wong-Baker FACES Pain Rating Scale (www.WongBakerFACES.org).

### 2.4. Language intervention

The language intervention protocol was based on studies that have successfully treated written language production. We adapted the basic design of a spell-study-spell procedure (Rapp and Glucroft 2009) to a lexical retrieval, oral and written naming paradigm (Beeson and Egnor 2006) to simultaneously target orthography, phonology and semantics. Although the intervention focused on written naming and spelling, this particular paradigm gave us the flexibility to accommodate the deficit in each variant (e.g., semantics in the svPPA, phonological paraphasias in lvPPA or apraxia of speech [AOS] errors in nfvPPA). The exact steps are described in previous publications of the overall trial results (Tsapkini et al. 2018). To evaluate whether therapy gains generalized in other words, the words of the untrained set were presented at all evaluation points.

### 2.5. Language and cognitive assessment: Verbal fluency

Participants were also evaluated with a series of standardized language and cognitive assessments. For the semantic fluency task, participants were instructed to name as many fruits, animals, and vegetables as possible, administered separately in the order listed here, in one minute per category (Benton et al. 1994). Scores used in the present analysis were calculated by adding the number of words generated in all three categories. Performance was assessed before, immediately after, two weeks after, and two months after each phase.

### 2.6. Imaging methods

Of the 36 participants, 29 had magnetic resonance imaging (MRI) scans—five were severely claustrophobic and two had pacemakers and were therefore excluded. MRI scans took place at the Kennedy Krieger Institute at Johns Hopkins University. Magnetization-prepared rapid acquisition gradient echo (MPRAGE) and resting-state functional MRI (rsfMRI) scans were acquired before treatment on a 3-Tesla Philips Achieva MRI scanner with a 32-channel head coil. T1-weighted MPRAGE sequence acquisition was performed according to the following parameters: a scan time of 6 minutes (150 slices); isotropic 1-mm voxel size; flip angle of 8°; SENSE acceleration factor of 2; TR/TE = 8/3.7 milliseconds (ms). Resting-state fMRI acquisition was performed according to the following parameters: scan time of 9 minutes (210 time-point acquisitions); slice thickness of 3 mm; in-plane resolution of 3.3×3.3 mm2; flip angle of 75°; SENSE acceleration factor of 2; SPIR for fat suppression; TR/TE = 2500/30 ms.

MPRAGE images were preprocessed and segmented into 283 regions of interest (ROIs) using MRICloud, a multi-atlas based, automated image parcellation approach. Preprocessing used a multi-atlas fusion label algorithm (MALF) and large deformation diffeomorphic metric mapping (LDDMM) (Tang et al. 2013; Mori et al. 2016), a highly accurate diffeomorphic algorithm that minimizes effects of atrophy or local space deformations on mapping. All images were processed in native space. Volumes for each ROI were normalized by total intracerebral volume (brain tissue excluding myelencephalon and cerebrospinal fluid) to control for relative regional atrophy.

Resting-state fMRI scans were preprocessed using MRICloud and included standard routines from the SPM connectivity toolbox for coregistration, motion, and slice timing correction; physiological nuisance correction using CompCor (Behzadi et al. 2007); and motion and intensity TR outlier rejection using “ART” (https://www.nitrc.org/projects/artifact_detect/). To correct for motion, ART detected “outlier” TRs (2 standard deviations for motion and 4 standard deviations for intensity), which were used in combination with the physiological nuisance matrix in the deconvolution regression for the remaining TRs.

Resting-state fMRI scans were co-registered with MPRAGE scans into the same anatomical space (native space); then 78 of the ROIs were parcellated on the rsfMRI scans. Average time courses for the voxels in each ROI were normalized, and correlations between ROI pairs were calculated and normalized with the Fisher z-transformation. Of the 78 ROIs, we chose the ones that comprise the language-network ROIs and are connected to the left IFG, the stimulated area (excluding the angular gyrus since it also belongs to the default mode network) as well as the right homologue of the stimulated area (the left IFG) since it connects directly to it. We, thus, included all the 78 pairs of the following 13 ROIs: the left and right pars opercularis, pars orbitalis, and pars triangularis of the inferior frontal gyrus (IFG_opercularis_L, IFG_opercularis_R, IFG_orbitalis_L, IFG_orbitalis_R, IFG_triangularis_L, IFG_triangularis_R), left middle temporal gyrus (MTG_L), left supramarginal gyrus (SMG_L), left superior temporal gyrus (STG_L), left inferior temporal gyrus (ITG_L), left fusiform gyrus (FuG_L), pole of the left middle temporal gyrus (MTG_L_pole) and pole of the left superior temporal gyrus (STG_L_pole).

### 2.7. Statistical analyses

#### 2.7.1. Evaluation of tDCS effects

We evaluated the effect of tDCS immediately after, at two weeks, and at two months post the intervention using the first-phase data only, in order to rid the estimation of any possible impact of carryover effects. In fact, we focus on the first-phase data throughout the statistical analysis for a similar reason, to eliminate possible treatment-phase interaction due to possibly insufficient wash-out period.

The additional tDCS effect compared to sham was evaluated as the average treatment effect (ATE), δ_(T vs S)_ = E[Y|T = 1] - E[Y|T = 0], where Y is the change in semantic fluency scores from baseline and T is the treatment assignment indicator (valued as 1 for tDCS, 0 for sham). We assumed that (Y_i_, T_i_), i = 1, …, n, is an independent and identically distributed (IID) sample, and that the actual observed Y_i_ is one of the two potential outcomes, Y_i_^T = 1^ and Y_i_^T = 0^, depending on the treatment assignment, i.e. Y_i_ = T_i_ Y_i_^T = 1^ + (1 - T_i_) Y_i_^T = 0^. These potential outcomes are defined as the changes would have been observed if the i-th subject had been assigned to tDCS or sham, was observed. Treatment randomization guarantees that the preassigned treatment variable, T_i_, is independent from the subsequent potential outcomes, Y_i_^T = 1^ and Y_i_^T = 0^, and that the assignment probability is constant and positive. The following baseline covariates were used to improve the efficiency of statistical tests: baseline semantic fluency (Lazar et al. 2010; DeMarco and Turkeltaub 2018), PPA variant, number of treatment sessions, sex, age, years post onset of symptoms, and total FTD-CDR severity and language severity measures.

The estimation of ATE was conducted using the Targeted Minimum Loss-Based Estimation (TMLE) method (van der Laan and Rose 2011). We chose the method mainly for its advantage of flexibility in selecting the covariates automatically (based on cross validations), which reduced subjective modeling choices of the researchers. Meanwhile, TMLE achieves the desired properties as other commonly practiced methods. For example, TMLE guarantees the estimation consistency and asymptotic normality in randomized trials as analysis of covariances (ANCOVA) estimators, as well as the doubly robust local efficiency as augmented inverse propensity score weighting (AIPW) estimators. The TMLE R package (Gruber and Laan 2012) has been made available, which can be directly applied for the aforementioned ATE estimation problem. The estimator configuration details and the simulation evidence supporting the choice of method are provided in the supplementary materials (Appendix A). Estimations of the ATE are reported as the effect sizes (Glass et al. 1981) adjusted by the first-phase before-after sham group standard deviation (SD= 3.00). The standard errors, Z-test statistics, p-values, and 95% confidence intervals are reported.

To handle missing observations in semantic fluency due to dropouts at two weeks post (two dropouts) and two months post (six dropouts), we assumed Missing at Random (MAR), that is, given the knowledge of the observed semantic fluency values and the series of observed baseline covariates, whether a subject happened to drop out was assumed to have no impact on the unobserved semantic fluency values. TMLE can be applied to handle such missingness in the outcome (van der Laan and Rose 2011), and can be directly implemented with the TMLE R package (Gruber and Laan 2012).

#### 2.7.2. Variant effects

Apart from the overall tDCS effect on semantic fluency and the functional connectivity pairs that predict it, we also investigated how it is modified by each variant subgroup at different time points. We assumed MAR and applied the linear mixed model (LMM) with random subject intercepts. In the fixed effect model, we included the following variables: the treatment assignment variable (tDCS and sham), the time point variable (after, 2 weeks and 2 months), as well as all the baseline covariates, were included as the main terms. Two-way interaction terms between time, treatment, and variant were added to the fixed effects. Three-way interaction terms were dropped in the final model, as none of these terms was significant with t-tests using Satterthwaite’s degrees of freedom (Kuznetsova et al. 2017), and these terms did not significantly improve the model fitting either in the likelihood ratio test (*χ*^2^(8) = 5.40, *p* = 0.714). Additional tDCS effects were evaluated at different time points (immediately after, two weeks post, and two months post) for each variant subgroup (nfvPPA, svPPA, lvPPA). Estimations of the effects (adjusted by the first-phase before-after sham group standard deviation), standard errors, Wald test statistics, p-values and 95% confidence intervals were reported.

#### 2.7.3. Prediction of Potentially Heterogeneous tDCS effects from non-imaging factors and baseline rsfMRI

We first investigated whether any of the non-imaging factors (baseline semantic fluency, PPA variant, number of treatment sessions, sex, age, years post onset of symptoms, and total FTD-CDR severity and language severity measures) was predictive of the individual tDCS effect. Secondly, we tested the imaging factors (correlations between the prespecified 13 language ROIs of the baseline rsfMRI; 78 ROI pairs in total). For the second task, to handle missingness in the baseline resting-state functional connectivity data, an inverse propensity score weighting (IPW) method was applied. Propensity scores were estimated using logistic regression with the imaging missingness and baseline covariates. The inverse propensity scores were then used as weights; each least square fitting using all participants in the non-imaging predictor selection procedure was replaced with weighted linear regression on the complete cases. The selection criteria based on LOOCV predictive R-squared increase for predicting pseudovalues remained the same. Below we present the details of our methods.

For each participant, the individual-specific additional tDCS effect, Y_i_^T = 1^ - Y_i_^T = 0^, which was defined as the difference between the potential change in semantic fluency had the participant been assigned to tDCS vs. had the individual been assigned to sham, may be heterogeneous. To evaluate which factors contributed to such inter-individual variability in the tDCS effect, we modeled them on the conditional average treatment effect (CATE), that is, E[Y_i_^T = 1^ - Y_i_^T = 0^ | X_i_] = E[Y|T = 1, X] - E[Y|T = 0, X], where X represents a group of predictive factors. An immediate challenge was that each individual was assigned to one of the two treatment groups; therefore, only one of the two potential scenarios was observed. To handle this issue, a nonparametric regression method using jackknife pseudovalues was applied (Efron and Tibshirani 1994; Rubin and van der Laan 2007; Tian et al. 2014). The jackknife pseudovalue, U_i_, of the *i*-th subject was calculated as a transformation of the observed behavioral outcome, U_i_ = 2Y_i_T_i_ – 2Y_i_(1 - T_i_); such pseudovalue maintains the same conditional expectation structure as directly modeling the CATE, i.e. E[U_i_ | X_i_ = x_i_] = E[Y | T = 1, X = x_i_] - E[Y | T = 0, X = x_i_]. Then the candidate factors were selected, depending on whether they were predictive to the resulted pseudovalues. Linearity was assumed and variable selection was conducted based on the leave-one-out cross validated (LOOCV) predictive R-squared. At each step of the forward selection, a threshold of 0.1 (10%) on the R-squared increase was applied to stop the selection procedure, otherwise the variable with the largest R-squared increase was selected. The predictive R-squared and the root mean squared error (RMSE) of each step, as well as the increase in predictive R-squared compared to the previous step, are reported for each round of the variable selection in Figure 3.

**Figure 2.**
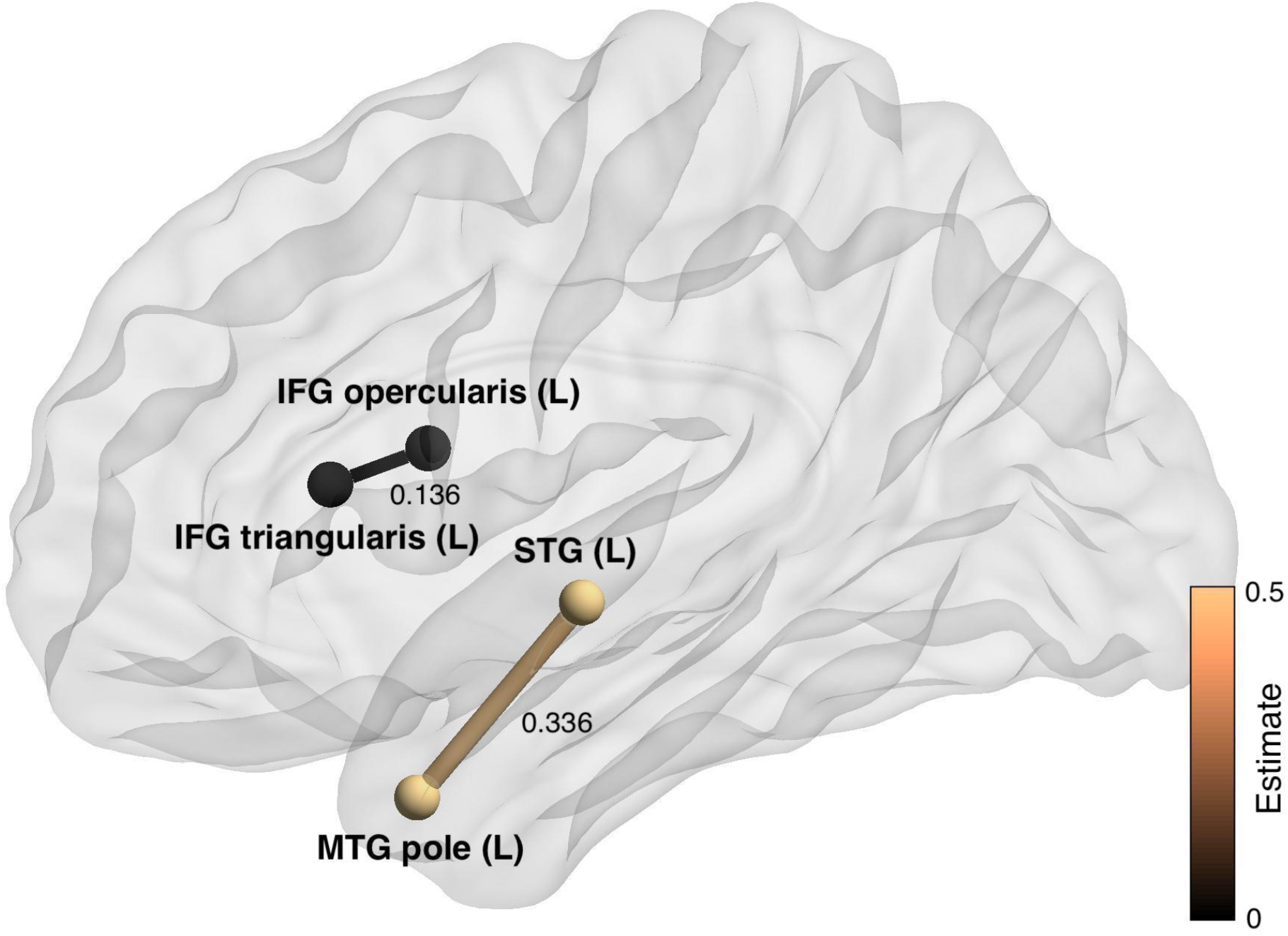
Visualization of the selected predictive imaging pairs for the additional tDCS effects. The positions of the nodes represent the average centers of each ROI from the cohort, rather than actual spatial distance. ROI pairs are plotted and connected if predictiveness of the baseline connectivity is confirmed. Thickness of the edge and the scale of the edge color represent on average how much additional semantic fluency increase one would expect for tDCS compared to sham, with 0.01 higher baseline Fisher z-transformed connectivity.

**Figure 3.**
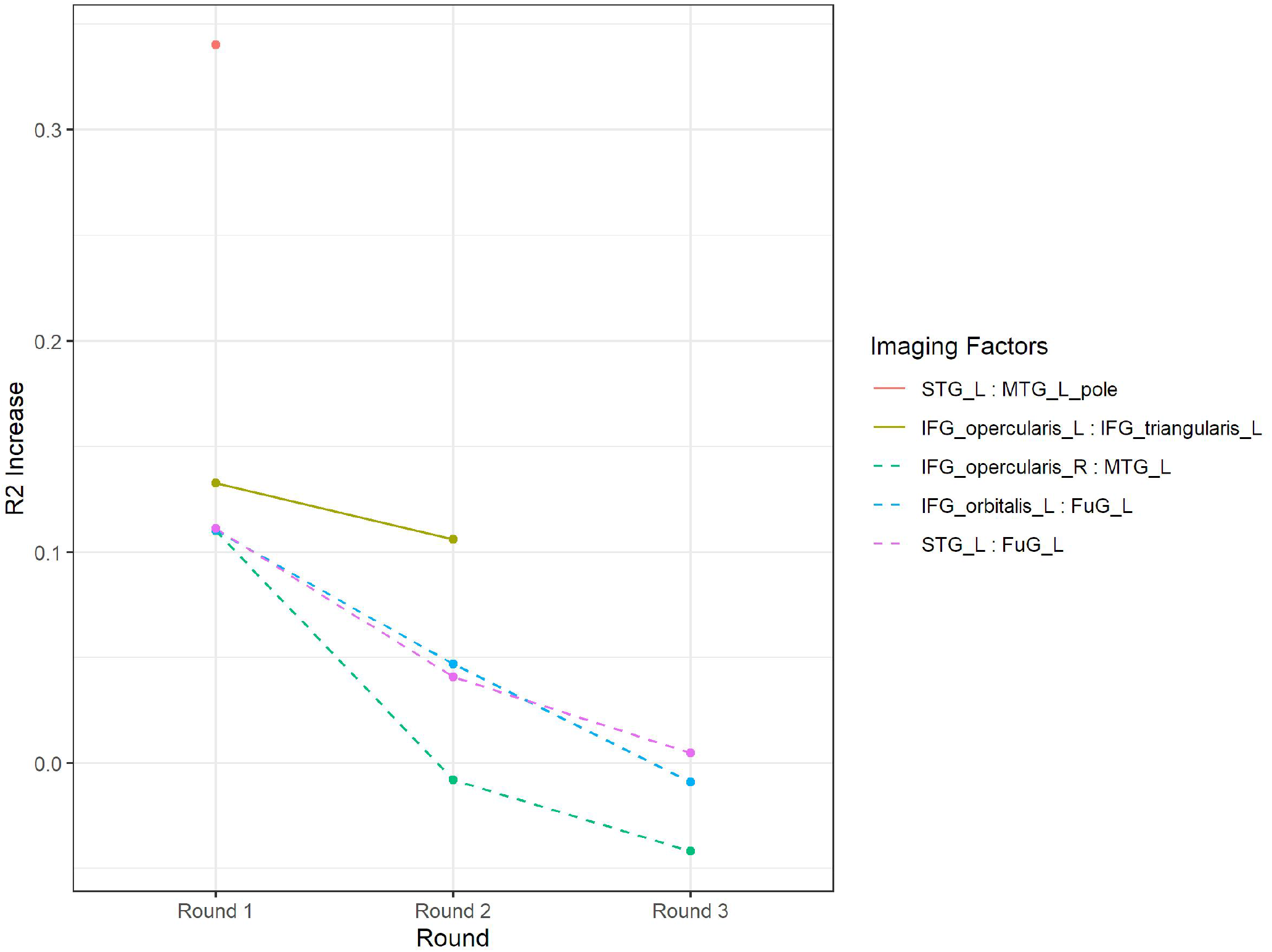
Functional connectivity pairs predicting the increases in R-squared for the individual tDCS effects. Solid lines represent the selected factors in each round of the regression, whereas dotted lines represent the factors that were not selected but also provided over 0.1 increase of predictive R-squared in the first round.

## 3. Results

### 3.1. tDCS tolerability

Some participants reported tingling, itching, or discomfort from the stimulation, but no episodes of intolerability or adverse effects occurred. The maximum reported Wong-Baker FACES pain rating scale for each daily session was averaged across sessions and participants, with a tDCS mean pain rating of 2.21 (standard deviation 2.48, range 0-10) and a sham mean rating of 2.14 (standard deviation 2.13, range 0-10).

### 3.2. tDCS effects on spelling accuracy

Previously, in the same patient cohort, we found that tDCS over the left IFG resulted in larger and longer-retained gains in letter accuracy for spelling trained items (words treated in therapy) and untrained items (words not treated in therapy as a measure of generalization to other items in the same task) as compared to sham (Tsapkini et al. 2014, 2018). This additional improvement with tDCS was retained up to 2 months. We refer to the previous study only as background information here. In the present paper, we extend these findings to semantic fluency as a measure of treatment generalization.

### 3.3. tDCS effects on semantic fluency

We present the additional tDCS effects (over sham) immediately after, two weeks and two months post-intervention from the first phase of intervention only, to avoid possible carryover effects. Sematic fluency scores ranged from 0-46 words. Scores were generated by the sum of fruits, animals, and vegetables generated in one minute each. All statistical analyses were performed the changes from the baseline in the summed scores of the three semantic categories.

#### 3.3.1. Evaluation of semantic fluency immediately after, two weeks post, and two months post the intervention

Restricting to the first-phase data, from before to after the intervention, the effect of tDCS vs. sham was significant, and the effect size was estimated as 1.35 (95% CI [0.30, 2.41], SE = 0.54, *Z* = 2.52, *p* = 0.012) adjusted for all the following covariates (baseline semantic fluency, PPA variant, number of treatment sessions, sex, age, years post onset of symptoms, and total FTD-CDR severity and language severity measures) as listed in Section *2.7.1*. Thus, tDCS showed an additional effect that is 1.35 times the standard deviation of the sham group’s change (SD=3) in the semantic fluency score.

Significant tDCS effect on semantic fluency was also confirmed two weeks post the intervention (effect size estimation 1.56, 95% CI: [0.40, 2.73], SE: 0.59, *Z* = 2.64, *p* = 0.008) but not two months post the intervention (effect size estimation 1.08, 95% CI: [-0.17, 2.33], SE: 0.64, *Z* = 1.69, *p* = 0.091).

#### 3.3.2. Variant effects

People with nfvPPA appeared to show the greatest generalization of improvement to semantic fluency, with the largest (compared to other variants) effect sizes at all 3 follow-up points: immediately after (estimated as 1.72), at two weeks post (1.71), and at two months post treatment (1.48). See Table 2. The effect size of tDCS in lvPPA was smaller than in nfvPPA immediately after (1.07), and at two weeks post (1.06) but diminished at two months post (0.83). The effect size of tDCS in svPPA was the smallest amongst variants at any follow-up point (0.55, 0.54 and 0.31, immediately after, at two weeks post and at two months post, respectively).

**Table 2.** Effect sizes for improvement in semantic fluency across variants immediately after, two weeks post, and two months post the intervention. The first phase data was used. Estimations were for the effect sizes divided by phase 1 before-after sham group standard deviation (SD= 3.00), adjusting for the baseline covariates as listed in Section *2.7.1*.

| | Estimate | SE | $\chi^2(1)$ | $p$ | Lower | Upper |
| --- | --- | --- | --- | --- | --- | --- |
| T vs S, nfvPPA, After | 1.72 | 0.58 | 8.71 | 0.003 | 0.58 | 2.86 |
| T vs S, lvPPA, After | 1.07 | 0.56 | 3.60 | 0.058 | -0.04 | 2.18 |
| T vs S, svPPA, After | 0.55 | 0.71 | 0.60 | 0.437 | -0.84 | 1.93 |
| T vs S, nfvPPA, 2week | 1.71 | 0.58 | 8.63 | 0.003 | 0.57 | 2.85 |
| T vs S, lvPPA, 2week | 1.06 | 0.58 | 3.31 | 0.069 | -0.08 | 2.20 |
| T vs S, svPPA, 2week | 0.54 | 0.70 | 0.59 | 0.442 | -0.84 | 1.91 |
| T vs S, nfvPPA, 2month | 1.48 | 0.62 | 5.62 | 0.018 | 0.26 | 2.70 |
| T vs S, lvPPA, 2month | 0.83 | 0.62 | 1.81 | 0.178 | -0.38 | 2.04 |
| T vs S, svPPA, 2month | 0.31 | 0.72 | 0.18 | 0.670 | -1.10 | 1.71 |

#### 3.3.3. Prediction of Potentially Heterogeneous tDCS effects: non-imaging factors

We tested the following non-imaging factors for predicting the additional tDCS effect on semantic fluency: baseline semantic fluency, PPA variant, number of treatment sessions, sex, age, years post onset of symptoms, and dementia and language severity (Frontotemporal Dementia Clinical Dementia Rating [FTD-CDR] overall sum and language measure, respectively) (Knopman et al. 2008) as we explain in the Statistical Analysis section (2.7.3). None of the non-imaging factors predicted the individual tDCS effect with R-squared increment thresholded at 0.1 (10%). However, dementia severity (overall FTD-CDR sum) and belonging to the nfvPPA group marginally predicted the additional tDCS effect in semantic fluency. Each provided 4.4% and 5.4% increases, respectively, in predictive R-squared and resulted in an accumulated R-squared of 9.8% (see Table 3).

**Table 3.**
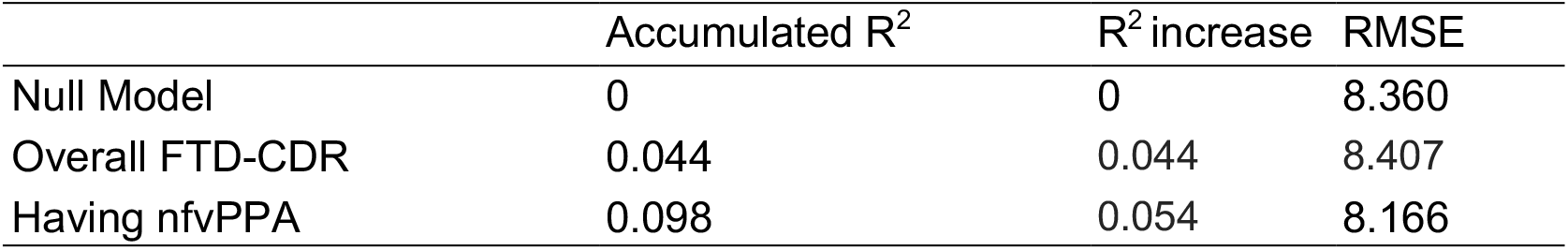
Non-imaging factors for individual tDCS effect prediction.

#### 3.3.4. Prediction of Potentially Heterogeneous tDCS effects: imaging factors

We tested a total of 78 resting-state functional connectivity pairs from 13 language network regions-of-interest (ROIs) as predictors for the additional tDCS effect on semantic fluency by taking into account statistically the individual heterogeneity of the participants. The whole analysis procedure is described in detail in Section 2.7.3. Since we had not found any single demographic or clinical factor to predict the additional tDCS effect above the R-squared minimum threshold of 10% (see above section 3.3.3), we did not enter any other factors in the regression model to save degrees of freedom given the sample size. Admittedly, the trade-off could be that the prediction effect would possibly be confounded by other baseline non-imaging factors. However, the comparison between the final models from the non-imaging and imaging predictors (Tables 3 and 4) in terms of the accumulated predictive R^2^ (0.098 vs 0.446) and the RMSE (8.166 vs 6.431) indicated that the imaging predictors outweighed the non-imaging ones in predictive value. Furthermore, the selected imaging factors are not significantly different in the three variant groups (Left STG : Left MTG pole, F(2, 21) = 0.13, p = 0.88; Left IFG opercularis : Left IFG triangularis, F(2, 21) = 0.76, p = 0.48; assuming missing completely at random) and are not significantly associated with Overall FTD-CDR (Left STG : Left MTG pole, Estimate = 8.58E-4, SD = 8.45E-3, t(22) = 0.10, p = 0.92; Left IFG opercularis : Left IFG triangularis, Estimate = −1.00E-2, SD = 1.37E-2, t(22) = − 0.73, p = 0.47; assuming missing completely at random); thus it was confirmed that their predictiveness for tDCS effects was not merely due to potential association with variant differences or dementia severity.

**Table 4.** Imaging factors that predicted the individual tDCS effect. Predictiveness was evaluated by the LOOCV (predictive) R^2^.

| | Accumulated $R^2$ | $R^2$ increase | RMSE |
| --- | --- | --- | --- |
| Null Model | 0 | 0 | 8.534 |
| Left STG : Left MTG pole | 0.340 | 0.340 | 7.250 |
| Left IFG opercularis : Left IFG triangularis | 0.446 | 0.106 | 6.641 |

Two baseline resting-state functional connectivity ROI pairs predicted the additional tDCS effect on semantic fluency above 10% of R-squared increase (0.1 threshold): (1) the left superior temporal gyrus (STG)-to-left medial temporal gyrus (MTG) pole and, (2) left IFG opercularis-to-left IFG triangularis (Table 4; Figure 2). The cumulative predictive R-squared of these two pairs was 0.446, i.e., 44.6% (RMSE=6.64). This result was confirmed in the refitted linear prediction model. Coefficients of this model were shown in Table 5. Higher baseline connectivity in these pairs was associated with higher additional tDCS effect. In addition, we monitored the changes of R^2^ increases in each round of variable selection (Figure 3). Note that in the first round three other imaging predictors provided R-squared increases greater than 0.1, but they were not selected because the Left STG: Left MTG pole had been selected for providing a larger R-squared increase.

**Table 5.** The prediction model for individual tDCS effects with the two selected imaging factors.

| | Estimate | SE | $t(21)$ | $P$ |
| --- | --- | --- | --- | --- |
| Intercept | -11.38 | 3.59 | -3.17 | 0.005 |
| Left STG : Left MTG pole | 33.60 | 7.11 | 4.72 | 0.000 |
| Left IFG opercularis : Left IFG triangularis | 13.59 | 4.79 | 2.84 | 0.010 |

## 4. Discussion

In the present study we evaluated the effects of anodal tDCS over left IFG (vs sham) on semantic fluency to provide causal evidence on the role of the left IFG in selective semantic retrieval. Previously, we (Tsapkini et al. 2018) and others (Cotelli et al. 2014; McConathey et al. 2017; Roncero et al. 2017) found positive effects of tDCS in PPA, especially in lexical retrieval tasks, such as oral and written naming following tDCS over the left IFG (Tsapkini et al. 2018). Given these results, we hypothesized that if the left IFG (triangularis, BA 45) subserves the function of selective semantic retrieval (Petrides 1995, 2014), particularly in selecting some aspect or subset of available information among competing alternatives (Thompson-Schill et al. 1997, 2005), then tDCS over this area will improve performance on a task of semantic selection, such as semantic verbal fluency, even if not explicitly trained. Given that selective semantic retrieval depends on functional (and likely structural) connections between the left IFG and temporal cortices (Tyler et al. 2011a; Margulies and Petrides 2013; Petrides 2014), we also hypothesized that baseline functional connectivity of these areas will predict the tDCS effect in semantic fluency. From a clinical perspective, we also examined the role of baseline functional connectivity between the stimulated area (left IFG) and its functionally connected areas as a biomarker for who would benefit more from tDCS. We found that performance in semantic fluency improved significantly more in the tDCS condition than in sham. The additional improvement was maintained up to two months post-treatment. In a subsequent analysis of which PPA variant may benefit more from tDCS, we found that this effect was stronger in nfvPPA. Resting-state functional connectivity between semantic representation areas and semantic control areas predicted the tDCS effect on semantic fluency: (1) the left MTG pole to left STG and (2) the left IFG pars opercularis to pars triangularis. Furthermore, higher baseline functional connectivity between the adjacent areas in these two pairs predicted larger increases in semantic fluency. Importantly, the effects were specific to selective semantic retrieval. In other words, there were no effects of tDCS on other cognitive tasks that do not depend on the left IFG, including digit span forward (on After-Before changes, 95% CI [-0.28, 0.89], Estimate = 0.31, SE = 0.29, *t*(34) = 1.06, *p* = 0.290) and Trail A (Tombaugh 2004) (on After-Before changes, 95% CI [-19.14, 28.89], Estimate = 4.87, SE = 11.19, *t*(14) = 0.44, *p* = 0.670), nor was there a tDCS effect of any index of general improvement, such as the Patient Health Questionnaire −9 [PHQ-9] (Kroenke et al. 2001) (on After-Before changes, 95% CI [-2.41, 5.41], Estimate = 1.50, SE = 1.82, *t*(14) = 0.82, *p* = 0.424). These findings demonstrate the specificity of positive far-transfer effects of tDCS to a non-trained task the depends on the left IFG, in the largest cohort of people with a neurodegenerative syndrome, PPA, to date. The present study significantly informs our neurocognitive understanding of the causal role of the left IFG in selective semantic retrieval, while also demonstrating important clinical implications on which patients will improve from tDCS over the left IFG.

The present study aligns with other tDCS studies in healthy aging and post-stroke aphasia that found tDCS to facilitate transfer of therapy effects to untrained tasks. For example, Marangolo et al. (2011) report that oral naming improved in post-stroke aphasia following apraxia of speech (AOS) training in conjunction with tDCS over the left IFG (Marangolo et al. 2011), and Meinzer et al. (2014) report that oral naming improved in mild cognitive impairment (MCI) after stimulation over the left motor cortex (M1)(Meinzer et al. 2014). With regard to tDCS studies in PPA, our study aligns with previous tDCS transfer effects on verbal fluency in smaller studies that targeted a variety of other language functions and tasks (Gervits et al. 2015; Roncero et al. 2017; Hosseini et al. 2019) ranging from generic tasks such as story-telling (Gervits et al. 2015) to oral naming (Roncero et al. 2017; Hosseini et al. 2019). The present study, thus, confirms in a large group of patients with adequate power that stimulation over the left IFG improves selective semantic retrieval even if not explicitly trained.

Another important finding in the present study is that the strength of functional connectivity between adjacent areas in both semantic processing areas (MTG and STG), as well as semantic control areas (IFG opercularis and triangularis), predicted the magnitude of tDCS effect on semantic fluency. This seems to be at odds with the atrophy in frontal areas, especially in nfvPPA who showed the largest tDCS effect. Given that we (Tao et al. under review) and others (Agosta et al. 2013; Mandelli et al. 2016) have not found a correlation between functional connectivity and atrophy, we would like to entertain the following possible hypotheses for the present results. (1) tDCS may be more beneficial on atrophied (but still viable) tissue, enhancing its lower baseline function. (2) Alternatively, tDCS may work better when other network or compensatory brain areas remain intact. People with nfvPPA, for example, have non-atrophied temporal regions (Gorno-Tempini et al., 2011). If functional connectivity within temporal regions best predicts the tDCS effects, perhaps their functioning, important for semantic storage and organization, compensates for frontal dysfunction and facilitates semantic retrieval. (3) Another possible hypothesis could be what we call ‘functional diaschisis’, i.e., compensatory brain regions (other IFG subregion or the right IFG) may mediate the response of the left IFG to tDCS. Despite the fact we have not found strong evidence for this hypothesis in the present study, the functional connectivity pairs in the different rounds of regression (shown in Figure 3) provide suggestions for the role of the right IFG and other temporal areas.

Lesion studies have shown that the neural substrates of semantic fluency are the left (or bilateral) IFG (particularly the IFG pars triangularis) and temporal areas, such as the MTG and ITG. More specifically, lesion studies have shown that the left temporal cortex stores information about semantic categories, and frontal areas are important in accessing this information (Mummery et al. 1999; Baldo et al. 2006). We also found that left ITG is associated with storage of lexical characteristics of both nouns and verbs in PPA (Race et al. 2013; Riello et al. 2018). With regard to verbal fluency in PPA, a previous study (Libon et al. 2009), as well as a recent one from our group (Riello et al. under review) where we controlled for dementia severity, found that atrophy in the anterior and inferior left temporal regions and right frontal regions were associated with semantic fluency.

Functional neuroimaging studies have provided strong evidence for the role of the left IFG in selective semantic retrieval from temporal areas (Thompson-Schill et al. 1997; Petrides 2006; Binder and Desai 2011; Rolheiser et al. 2011a; Tyler et al. 2011a). In a systematic review of fMRI studies related to semantic fluency, Costafreda and colleagues (2006) supported the association between semantic fluency and brain activation (BOLD signal) in the anterior and ventral portions of the left IFG (Costafreda et al. 2006). Thompson-Schill and colleagues argued that the left IFG triangularis is important for the selection of some aspect or subset of available information among competing alternatives, e.g., semantic category (Thompson-Schill, Aguirre, D’Esposito, & Farah, 1999; Thompson-Schill et al., 1997). Many studies have implicated the left IFG in strategic lexical retrieval pointing to the differential roles of the subdivided left IFG: the pars opercularis (BA 44) for phonologically-cued implicated retrieval and the pars triangularis (BA 45) for strategic semantically-cued retrieval (implicating BA 45) (Katzev et al. 2013; Petrides 2014). In particular, Amunts and colleagues (Amunts et al. 1999, 2004; Gurd et al. 2002) combining evidence from fMRI and cytoarchitectonic mapping, showed that although both BA 44 and BA 45 participate in verbal fluency, they do so differently: BA 44 is most likely involved in high-level programming of speech production, while BA 45 is more involved in semantic aspects of language processing (Grodzinsky and Amunts 2006). The present study adds causal evidence from tDCS for the role of the left IFG in selective semantic retrieval of verbal information stored in long-term memory in the temporal lobe (Petrides 1995; Thompson-Schill et al. 1997, 2005).

Interestingly, in the present study the two most significant functional connectivity pairs that predicted the tDCS effect on semantic fluency correspond to areas at the edges of the extreme capsule fasciculus, a white-matter bundle connecting the left IFG triangularis to temporal areas as part of the ventral language stream (Hickok and Poeppel 2007; Saur, Kreher, Schnell, Kümmerer, Kellmeyer, Vry, Umarova, Musso, Glauche, Abel, et al. 2008; Friederici 2009). Recent tractography studies using sensitive DTI in humans have traced this fasciculus (Frey et al. 2008) and demonstrated how it connects the IFG with anterior temporal and superior temporal regions. Furthermore, several studies have shown that the extreme capsule is important for semantic processing and comprehension (Saur, Kreher, Schnell, Kümmerer, Kellmeyer, Vry, Umarova, Musso, Glauche, and Abel 2008; Friederici 2009; Tyler et al. 2011b). Although this bundle is sometimes hard to detect and many times considered as part of the uncinate, in vivo tracing studies in humans and the macaque have shown that the uncinate connects prefrontal regions to anterior temporal and involves emotional regulation (Petrides and Pandya 2009). The structural white matter connections between semantic control (the left IFG) and semantic processing areas (the anterior, inferior, and middle temporal and fusiform gyri) may allow the left IFG to act as the neural substrate of selective retrieval of categorical information stored in temporal areas. In a recent study, we have also identified the white-matter integrity of this bundle as a significant predictor of tDCS but not language therapy alone (sham) for trained words during written naming therapy (Zhao et al. under review). However, in that study the contribution of the structural connectivity to the tDCS effect was much more modest (12%) than here (46%) showing that functional connectivity is a better predictor of the tDCS effects in PPA.

Functional connectivity has also been identified as a mechanism of tDCS effects in previous studies by Meinzer and colleagues for healthy aging (Meinzer et al. 2012, 2013) and MCI (Meinzer et al. 2014), as well as by our group for PPA (Ficek et al. 2018). These studies identified decreases of functional connectivity between long-distance areas belonging to the same network (between the stimulated left IFG and temporal areas) as a tDCS mechanism. How do these previous decreases between distal areas as a tDCS mechanism reconcile with the present increases in correlation between adjacent areas as a predictor for tDCS effects? Previous studies in neurodegenerative disorders, including AD (Dickerson et al. 2005; Bakker et al. 2012) and PPA (Tao et al. under review; Mandelli et al. 2016), have identified increases of functional connectivity in areas that are at risk of atrophy and are subsequently lost as functional hubs. These increases have been interpreted as manifestations of a probable compensatory mechanism (Dickerson et al. 2005; Meinzer et al. 2012, 2014) or as manifestations of the disease progression itself since they did not always correspond to sustained performance (Bakker et al. 2012). Meinzer and colleagues (2013) suggested that tDCS downregulates that hyperactivity or hyperconnectivity (Meinzer et al. 2013), and our previous findings converged to this suggestion (Ficek et al. 2018).

The present tDCS study provides causal evidence that the left IFG is a critical area for selective semantic retrieval even in neurodegenerative conditions such as PPA. Furthermore, stronger functional connectivity within semantic processing and semantic control areas predict better performance in a selective semantic retrieval task (semantic fluency). The results of the present study have also important clinical implications, indicating that nfvPPA may benefit more from tDCS over the left IFG. Furthermore, baseline functional connectivity in semantic processing and semantic control areas may be biomarkers for selecting patients who will benefit more from tDCS. Further work confirming mechanisms by which tDCS may affect functional connectivity or be influenced by fronto-temporal structural or functional connectivity is warranted.

## Data Availability

We will make the data and associated documentation available to users only under a data-sharing agreement that provides for: (1) a commitment to using the data only for research purposes and not to identify any individual participant; (2) a commitment to securing the data using appropriate computer technology; (3) proper acknowledgment of data source; and (4) a commitment to destroying or returning the data after analyses are completed.

## Acknowledgements

We would like to thank our participants and referring physicians for their dedication and interest in our study. Funding: This work was supported by grants from the Science of Learning Institute at Johns Hopkins University and by the National Institutes of Health (National Institute of Deafness and Communication Disorders) through award R01 DC014475 to KT. AH was supported by NIH (NIDCD) through awards R01 DC03681, R01 DC011317 and P50 DC014664.

